# Cognitive Flexibility is Critical for Postural Control in People with Parkinson Disease

**DOI:** 10.1101/2025.09.26.25336649

**Authors:** Matt Leedom, Patti Berg-Poppe, Arturo I. Espinoza, Arun Singh

## Abstract

**Background:** Postusral instability is a common and debilitating symptom in Parkinson disease (PD) that impacts daily functioning and quality of life. Executive function, a domain of cognitive functioning, has been associated with postural control. Impairments in executive function, including cognitive flexibility, may lead to postural instability in individuals with PD. This study aimed to examine the relationship between postural instability and cognitive flexibility in PD. While broader executive function domains were explored, cognitive flexibility was hypothesized to be particularly relevant to postural instability. Cognitive assessments were selected to specifically evaluate this domain alongside other key aspects of executive function.

**Objective:** To examine the relationship between postural instability and cognitive flexibility in PD.

**Methods:** Thirty-six participants, including 12 people with PD who had postural instability (PDPI+), 12 people with PD without postural instability (PDPI–), and 12 age-matched healthy controls participated. Cognitive assessments were performed using the MoCA and the NIH Toolbox-Cognition Battery. A Clinical Balance Systems (CBS) score was also calculated during clinical evaluation.

**Results and Discussion:** Data analysis included ANOVA, ANCOVA, Pearson correlation, and partial correlation analyses. The PDPI+ group exhibited significant cognitive deficits compared to healthy controls, particularly in cognitive flexibility. Cognitive flexibility emerged as a critical factor linked to postural instability. In all participants who had PD, strong negative correlations were observed between CBS scores and cognitive flexibility, suggesting a robust connection between cognitive flexibility and postural control. These associations remained significant even after controlling for age, disease duration, and levodopa equivalent daily dose.

**Conclusions:** Our findings underscore the significant relationship between cognitive flexibility and postural control in individuals with PD. The results highlight the potential role of targeted interventions to improve cognitive flexibility and, consequently, postural stability in people with PD who have postural instability.

## INTRODUCTION

Postural control is essential for daily activities and is often impaired in Parkinson disease (PD), leading to falls. Almost 60% of people with PD experience at least one fall, and 40% of people with PD have frequent falls in relation to symptoms of postural control (Pelicioni et al, 2019). Executive function, a cognitive domain that is highly associated with postural control, refers to a set of cognitive processes involved in planning, organizing, initiating, and monitoring goal-directed behaviors (Buated et al, 2016; Chaudhary et al, 2020). This includes abilities such as attention, working memory, inhibitory control, and cognitive flexibility. Impairments in executive function, particularly in aspects like attention, can significantly contribute to difficulties in maintaining balance and postural stability (Amboni et al, 2013). Furthermore, previous research suggests a relationship between cognitive dysfunction and motor symptoms of postural instability/gait disturbance (PIGD), which could implicate distinct underlying neural pathways (Kelly et al, 2015).

As people with PD experience disease progression, deficits in executive function substantially impact ability of patients to make rapid postural adjustments and coordinate movements, leading to increased postural instability and gait dysfunction (Barboza et al, 2023; Fernandes et al, 2016; Morris et al, 2019). While executive function is strongly associated with postural control, it is important to recognize that attention, as a component of executive function, plays a pivotal role across various cognitive domains involved in maintaining balance and posture (Amboniet al, 2013). This includes visuospatial processing and motor planning, highlighting the interconnectedness of cognitive processes in the complex task of postural control.

Deficits in global cognitive function and executive function have been associated with more severe impairments in posture, balance, and gait symptoms in PD (Kellyet al, 2015; Pal et al, 2016; Sousa et al, 2021). Our preliminary report has also highlighted a robust relationship between the deterioration of postural control and midfrontal cortical oscillations linked to cognitive processing within this population (Bosch et al, 2021). These findings underscore the crucial involvement of cognitive networks in postural control, emphasizing the intricate interactions between multiple cognitive processes (Johansson et al, 2019; Kellyet al, 2015; Landers et al, 2005; Santamato et al, 2015).

Comprehensive studies examining the specific effects of different cognitive domains on postural control in PD, especially among individuals experiencing instability, are currently limited. Previous research has suggested significant connections between global cognitive deficits, executive function deficits, and PIGD symptoms (Kellyet al, 2015). However, contrasting findings show mixed evidence for the relationships between PIGD symptoms and specific cognitive components (Green et al, 2002; Kellyet al, 2015; Lyros et al, 2008; Palet al, 2016; Sousaet al, 2021). In this study, we sought to build on previous knowledge and address remaining gaps by conducting a comprehensive assessment of cognitive functions, using additional measures of specific cognitive domains, in individuals with Parkinson disease exhibiting postural instability (PDPI+), those without postural instability (PDPI–), and age-matched healthy control participants. Our primary objective was to investigate whether cognitive flexibility is specifically associated with postural instability in PD. Additionally, we examined broader executive function domains to determine their potential contributions. To align with this objective, we selected cognitive assessments that provide a targeted evaluation of cognitive flexibility while also capturing other aspects of executive function. The NIH Toolbox Cognition Battery (NIHTB-CB) includes validated measures of cognitive flexibility and executive processing, allowing for a focused yet comprehensive assessment of the constructs relevant to our hypothesis (Nasreddine et al, 2005; Weintraub et al, 2013). Another tool that was selected, the Montreal Cognitive Assessment (MoCA), is a widely recognized cognitive screening tool that assesses several cognitive domains, including executive functions, visuospatial functions, naming, memory, attention, language, abstraction, delayed recall and orientation. By employing these assessment tools, we aimed to delineate the contribution of various cognitive functions, including cognitive flexibility, to postural control in people with PD.

Cognitive flexibility is the ability to adapt behavior to changing environments (Dajani and Uddin, 2015; Diamond, 2013; Hanes et al, 1995). This includes stopping one task, preparing new responses, and continuing the task with these adjustments. Processes involved in cognitive flexibility include salience detection and attention, working memory, inhibition, and shifting (reconfiguring a response set) (Dajani and Uddin, 2015). The highly distributed neural network responsible for cognitive flexibility includes regions within the frontoparietal cortex (e.g., cortical association areas, premotor cortex, inferior and superior parietal cortices), as well as additional areas such as the inferior temporal cortex, occipital cortex, and subcortical structures including the caudate and thalamus (Dajani and Uddin, 2015). Many of the same structures responsible for cognitive flexibility are frequently compromised in PD, particularly as the disease advances. Given the overlap of brain structures involved in both cognitive flexibility and cognitive control deficits in people with PD, we hypothesized that, when compared to other cognitive domains, cognitive flexibility serves as a predominant cognitive domain correlating with postural instability in PD.

## METHODS

### Participants

Thirty-six individuals (12 PDPI+, 12 PDPI–, and 12 age-matched healthy controls) were recruited for this study. Participants with PD included a total of 17 males and 7 females. Control participants included 6 males and 6 females. All participants were of Caucasian ethnicity. Participants adhered to the protocols approved by the institutional review boards of the University of Iowa and the University of South Dakota, including the completion of informed consent procedures. Clinical demographics are shown in Table 1. Participants with PD were asked to continue their levodopa regimen, as regularly prescribed, to mimic real-world conditions and avoid off-medication-related complications, such as falls (Frenklach et al, 2009; Landerset al, 2005; Nutt et al, 2011; Pelicioniet al, 2019). Potential participants with tremor and levodopa-induced dyskinesia and those who were wheelchair-dependent were excluded from the study. The NIHTB-CB was completed using a tablet. Participants experiencing tremor and dyskinesia can face challenges in performing fine motor tasks on a tablet, thus necessitating their exclusion. To account for medication effects, a validated calculator was used to determine the levodopa equivalent daily dose (LEDD) (Nyholm and Jost, 2021). LEDD was subsequently utilized as one of the covariates in statistical analyses.

**Table 1.** Demographics and clinical characteristic measurements.

|  | **Control**<br>**(n = 12)** | **PD**<br>**(n = 24)** | **PDPI–**<br>**(n = 12)** | **PDPI+**<br>**(n = 12)** | **Control vs.**<br>**PD**<br>**(n=36)** | **PDPI– vs. PDPI+**<br>**(n=24)** |
| --- | --- | --- | --- | --- | --- | --- |
|  |  |  |  |  | **Independent**<br>**t-test** | **Independent**<br>**t-test** |
| Age<br>(years) | 69.5 ± 9.4 | 68.5 ± 7.8 | 65.5 ± 8.4 | 71.5 ± 6.2 | 0.34 (p = 0.74) | -2.00 (p = 0.06) |
| mUPDRS | — | — | 10.8 ± 6.0 | 19.8 ± 4.7 | — | -4.10 (p < 0.001)** |
| DD<br>(years) | — | — | 5.8 ± 6.6 | 7.8 ± 4.6 | — | -0.83 (p = 0.42) |
| LEDD<br>(mg) | — | — | 739.6 ± 326.6 | 1068.3 ± 450.8 | — | -2.05 (p = 0.05) |

mUPDRS, motor Unified Parkinson's Disease Rating Scale; DD, Disease duration; LEDD, Levodopa equivalent daily dose.
Statistics presented as t-value (p-value). \*\*p < 0.001.

Participants for the PDPI+ group were identified based on their score on item 3.12, Postural Stability, of the Movement Disorder Society Unified Parkinson Disease Rating Scale (MDS-UPDRS) Part III, Motor Examination subscale. They were placed into the PDPI+ group if this score exceeded 0 (max. 4). This classification was confirmed by a movement disorder expert. Additionally, a clinical balance score (CBS), calculated as the sum of MDS-UPDRS items 3.8, 3.9, 3.12, and 3.13 (max. 16 = + Leg Agility, + Arising from Chair, + Postural Stability, + Posture) was computed for all participants with PD (Boschet al, 2021; Perera et al, 2018).

### Clinical Assessments

Several clinical assessments were used to measure domains of cognitive functioning as well as disease severity. The MDS-UPDRS was used to determine the severity of disease in each of the participants with PD. To measure cognition function, the MoCA and NIHTB-CB tests were administered. The MoCA includes items that measure short-term memory; visuospatial abilities; executive functions; attention, concentration, and working memory; language; and orientation to time and place. The cognitive tasks used from the NIHTB-CB included: the Dimensional Change Card Sort (DCCS) for cognitive flexibility, the Flanker Inhibitory Control and Attention (FICA) task for inhibitory control, Pattern Comparison Processing Speed (PCPS) for processing speed, Picture Sequence Memory (PSM) for episodic memory, and Picture Vocabulary (PV) test for language skills. These assessments were conducted in a laboratory setting.

The MoCA and NIHTB-CB are both widely used and validated measures of cognitive function. The MoCA has demonstrated good reliability and validity for detecting global cognitive impairment in PD populations. The NIHTB-CB is a comprehensive battery of cognitive tests that has been validated for use in various populations, including PD. In our study, all assessments, including the MoCA and NIHTB-CB, were conducted in the morning session after participants had taken their morning medications.

### Data Analyses

The obtained cognitive test scores were converted into fully corrected T-scores, accounting for age and other demographic variables (education, gender, and race/ethnicity). All statistical analyses were performed using the Matlab statistical toolbox. We conducted Kolmogorov-Smirnov (KS) tests on the DCCS of both control and PD data to assess normality. The results indicated that the control data (p = 0.29) and PD data (p = 0.68) were normally distributed.

To compare cognitive scores between healthy controls, PDPI–, and PDPI+ groups, one-way analysis of variance (ANOVA) tests were performed. These were then followed by pairwise comparisons with Bonferroni corrections. For further analysis, Pearson correlation tests were utilized to explore associations between cognitive scores in all participants with PD. To account for multiple correlations, p-values were adjusted using Bonferroni corrections.

To assess the differences between PDPI+ and PDPI– groups, and to investigate the relationship between cognitive measures and CBS in all participants with PD, analysis of covariance (ANCOVA) tests and Pearson partial correlations were conducted. Covariates, including age, disease duration (DD), and Levodopa equivalent daily dose (LEDD), were introduced sequentially—first individually, then age and DD combined, and finally, all three covariates (age, DD, and LEDD) simultaneously. Significance levels were presented both with uncorrected (p = 0.05) and Bonferroni-corrected (corrected p = 0.008) p-values to address the multiple correlation tests conducted in this study.

## RESULTS

### Clinical Characteristics Measurements

Detailed statistical comparisons are summarized in Table 1. When contrasting the demographic and clinical characteristics between groups, few significant findings emerged. There were no statistically significant differences in age between the healthy controls and people with PD (p = 0.74). When comparing PDPI-and PDPI+ groups, neither age nor disease duration (DD) displayed significant differences either; however, age showed a trend toward significance (p=0.06), whereas DD did not (p=0.42). A comparison between PDPI– and PDPI+ groups did reveal significant differences in MDS-UPDRS scores (p < 0.001), indicating variations in motor symptoms.

Additionally, there was a borderline significant difference in LEDD (p = 0.05), indicating variations in medication usage.

### ANOVAs: Cognitive Differences Among Controls and PD Groups

To comprehensively compare the cognitive performance of control participants, PDPI–, and PDPI+ groups, pairwise comparisons were performed (Fig. 1). An adjusted p-value of 0.017 was utilized, based on Bonferroni correction, to ensure the robustness of findings.

**Fig. 1.**
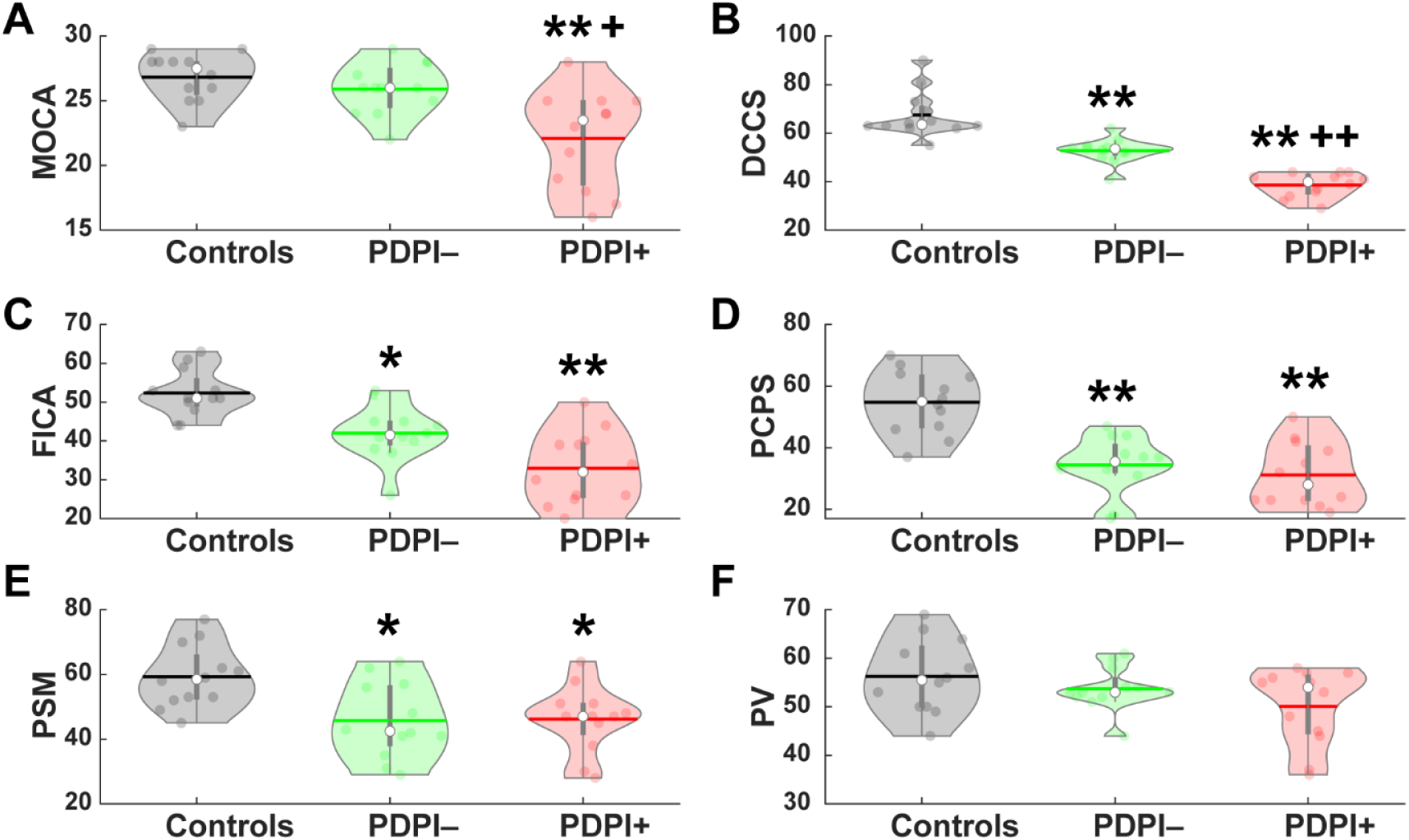
Cognitive performance scores of healthy controls, PDPI–, and PDPI+ participants. (A) MoCA scores, (B) DCCS scores, (C) FICA scores, (D) PCPS scores, (E) PSM scores, and (F) PV scores. Statistical significance denoted as corrected *p<0.017 vs. controls, **p<0.001 vs. controls, +p<0.017 vs. PDPI–, ++p<0.001 vs. PDPI–. MoCA, Montreal Cognitive Assessment; DCCS, NIHTB-CB Dimensional Change Card Sort Test; FICA, Flanker Inhibitory Control and Attention Test; PSM, Picture Sequence Memory Test; PCPS, Pattern Comparison Processing Speed test; PV, Picture Vocabulary test.

When evaluating MoCA scores, significant differences between the groups were identified (F(2,33) = 10.5, p = 0.001). Specifically, while there were no meaningful differences when comparing the PDPI– group to healthy control participants (p = 1), the PDPI+ group displayed a significant decline in MoCA scores relative to controls (p = 0.001) and PDPI– group (p = 0.004).

When assessing the DCCS test, a measure of cognitive flexibility, significant variations were observed among the groups (F(2,33) = 52.3, p = 0.001). Control participants exhibited significantly better performance on this test compared to both PDPI– (p = 0.001) and PDPI+ (p = 0.001) groups. Moreover, a significant difference was also noted between PDPI– and PDPI+ groups (p = 0.001). This finding highlights meaningful differences in cognitive flexibility performance in both PD groups, compared to health controls, and significant differences among people with PD who have postural instability and those who do not.

Group differences were also observed on the FICA test (F(2,33) = 19.3, p = 0.001). Control participants displayed a significant difference in cognitive performance compared to PDPI– (p = 0.007) and PDPI+ (p = 0.001) groups. No significant differences emerged between PDPI– and PDPI+ groups (p = 0.02).

When assessing the performance on the PCPS test, significant group differences were again observed (F(2,33) = 19.45, p = 0.001). Control participants displayed a performance that was significantly better than both PDPI– (p = 0.001) and PDPI+ (p = 0.001) groups. However, no significant differences were observed between PDPI– and PDPI+ groups (p = 1) in this domain.

On the PSM assessment, group differences were again evident (F(2,33) = 6.26, p = 0.005). Control participants demonstrated significantly better performance on this test compared to PDPI– (p = 0.011) and PDPI+ (p = 0.015) groups. However, no significant difference emerged between PDPI– and PDPI+ groups (p = 1) in relation to episodic memory, as evidenced by this test.

Lastly, on the PV test, group differences were not statistically significant (F(2,33) = 2.47, p = 0.1). There were no significant differences between control participants and either PDPI– (p = 1) or PDPI+ (p = 0.1) groups. Moreover, there were no statistically significant differences in performance between the PDPI– and PDPI+ groups (p = 0.59).

### ANCOVAs: Cognitive Differences Between PDPI– and PDPI+

In order to compare cognitive measures between PDPI– and PDPI+ groups, ANCOVAs were also performed. Several covariates were controlled for, including age, DD, and LEDD. Detailed statistical outcomes of ANCOVAs are demonstrated in Table 2. The data revealed significant findings based on the results of several cognitive measures.

**Table 2.** ANCOVAs between PDPI-vs PDPI+: Controlling for Age, Disease Duration (DD), Levodopa Equivalent Daily Dose (LEDD).

| Test | Age (Years) | DD | LEDD | Age & DD | Age, DD, & LEDD |
| --- | --- | --- | --- | --- | --- |
| MOCA | F(1,21) = 6.51<br>(p = 0.019)* | F(1,21) = 8.33<br>(p = 0.009)** | F(1,21) = 5.29<br>(p = 0.03)* | F(1,20) = 5.34<br>(p = 0.03)* | F(1,19) = 2.42<br>(p = 0.14) |
| DCCS | F(1,21) = 36.37<br>(p < 0.001)*** | F(1,21) = 49.15<br>(p < 0.001)*** | F(1,21) = 40.40<br>(p < 0.001)*** | F(1,20) = 37.99<br>(p < 0.001)*** | F(1,19) = 29.78<br>(p < 0.001)*** |
| FICA | F(1,21) = 6.04<br>(p = 0.02)* | F(1,21) = 6.12<br>(p = 0.02)* | F(1,21) = 5.27<br>(p = 0.03)* | F(1,20) = 5.12<br>(p = 0.04)* | F(1,19) = 4.13<br>(p = 0.06) |
| PSM | F(1,21) = 0.02<br>(p = 0.89) | F(1,21) = 0.01<br>(p = 0.92) | F(1,21) = 0.10<br>(p = 0.76) | F(1,20) = 0.02<br>(p = 0.89) | F(1,19) = 0.14<br>(p = 0.71) |
| PCPS | F(1,21) = 0.45<br>(p = 0.51) | F(1,21) = 0.64<br>(p = 0.43) | F(1,21) = 0.28<br>(p = 0.61) | F(1,20) = 0.45<br>(p = 0.51) | F(1,19) = 0.12<br>(p = 0.73) |
| PV | F(1,21) = 1.21<br>(p = 0.28) | F(1,21) = 1.42<br>(p = 0.25) | F(1,21) = 0.45<br>(p = 0.51) | F(1,20) = 0.74<br>(p = 0.40) | F(1,19) = 0.06<br>(p = 0.82) |

MOCA, Montreal Cognitive Assessment; DCCS, NIHTB-CB Dimensional Change Card Sort Test; FICA, Flanker Inhibitory Control and Attention Test; PSM, Picture Sequence Memory Test; PCPS, Pattern Comparison Processing Speed test; PV, Picture Vocabulary test.
Statistics presented as t-value (p-value). \*p < 0.05; \*\* p < 0.01; \*\*\* p < 0.001.

Related to the MoCA, significant differences between groups were observed in multiple comparisons. There was a significant effect of age on MoCA scores (F(1,21) = 6.51, p = 0.019), suggesting that age differences might influence cognitive performance measured by the MoCA. Disease duration (DD) also significantly impacted MoCA scores (F(1,21) = 8.33, p = 0.009), suggesting differences in cognitive function related to the duration of the disease. Additionally,

LEDD showed statistically significant effect on performance (F(1,21) = 5.29, p = 0.03), indicating a relationship between medication dosage and cognitive outcomes. When controlling for both age and DD (F(1,20) = 5.34, p = 0.03), the effect remained significant. However, when controlling for all three covariates (age, DD, and LEDD), the effect was not significant (F(1,19) = 2.42, p = 0.14). On the DCCS test, all models showed highly significant effects (p < 0.001). When controlling for age (F(1,21) = 36.37, p < 0.001), DD (F(1,21) = 49.15, p < 0.001), and LEDD (F(1,21) = 40.40, p < 0.001) individually, performance on the DCCS test indicated significant differences between PDPI-and PDPI+ groups under each condition. When controlling for age and DD together (F(1,20) = 37.99, p < 0.001), the differences in cognitive flexibility remained significant. These results revealed a consistent association between these covariates and performance on the DCCS test amongst people with PD. Additionally, in contrast to findings for the MoCA, a significant difference between the PDPI– and PDPI+ groups in performance on the DCCS test, while controlling for all three covariates, including age, DD, and LEDD, was observed (F(1,19) = 29.78, p < 0.001).

Similar to the MoCA, all models examining performance on the FICA, except the one with all three covariates, revealed significant effects. When assessing group differences in performance of inhibitory control, the significance level decreased when more covariates were included. The effects of age (F(1,21) = 6.04, p = 0.02), DD (F(1,21) = 6.12, p = 0.02), and LEDD (F(1,21) = 5.27, p = 0.03) each individually exhibited statistical significance within this cognitive domain. When controlling for age and DD (F(1,20) = 5.12, p = 0.04) together, statistical significance persisted. However, when all three covariates (age, DD, and LEDD) were controlled simultaneously (F(1,19) = 4.13, p = 0.06), the effect did not reach statistical significance, although there was a trend towards significance.

In contrast, neither the PCPS nor the PSM tests demonstrated significant effect on cognitive performance in any of the comparisons (all p > 0.05). These results suggest that performance on these tests is not influenced by age, DD, LEDD, or their combinations, and sheds light on the intricate interplay between clinical factors and cognitive performance in people with PD.

### CBS Correlations with Cognitive Measures

To examine the relationship between cognitive measures and Clinical Balance Systems (CBS) score, correlation analyses were performed (Fig. 2A). The MoCA exhibited a significant negative correlation (p = 0.005) and the DCCS test showed a robust negative correlation with CBS scores (p = 0.001). This strong association suggested that impaired cognitive flexibility, as assessed by the DCCS test, was closely related to the severity of CBS symptoms in people with PD. The FICA test also displayed a significant negative correlation with CBS scores (p = 0.025). However, no statistically significant correlations were observed between CBS scores and PCPS, PSM, or PV tests. Furthermore, upon employing a more stringent Bonferroni-corrected p-value threshold of 0.008 to mitigate the possibility of false-positive findings, significant correlations between CBS scores and both the MoCA and the DCCS test were identified. These findings contribute to a deeper understanding of the cognitive factors influencing the manifestation of CBS in PD.

**Fig. 2.**
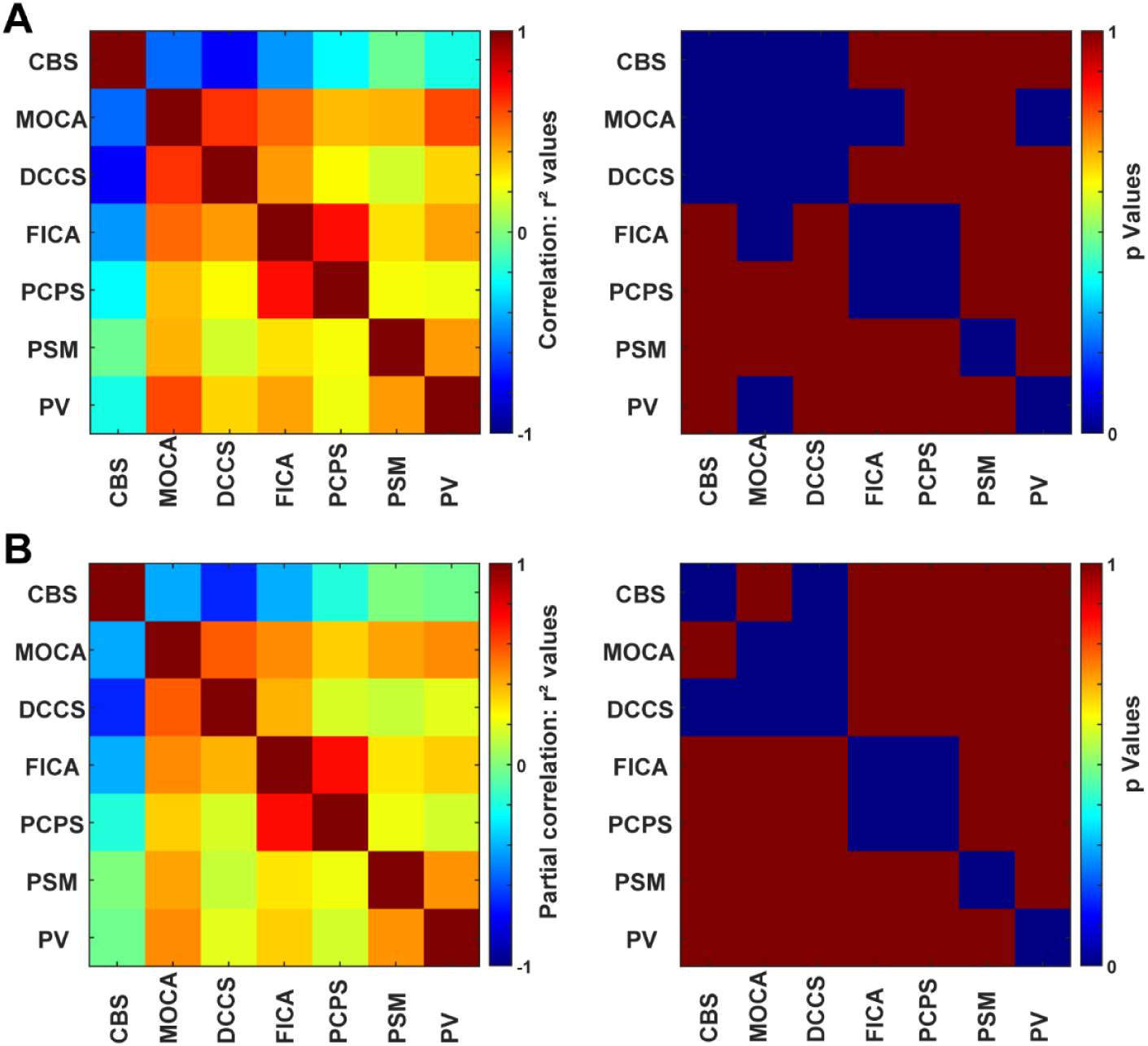
Correlation and Partial Correlation Analyses. A) Correlation coefficient and p-value matrices. B) Partial correlation coefficient and p-value matrices, controlling for age, disease duration, and LEDD. To enhance visualization, p-values exceeding 0.008 (corrected) were set to 1, and p-values below 0.008 were set to 0.001. CBS, clinical balance score; MoCA, Montreal Cognitive Assessment; DCCS, NIHTB-CB Dimensional Change Card Sort Test; FICA, Flanker Inhibitory Control and Attention Test; PSM, Picture Sequence Memory Test; PCPS, Pattern Comparison Processing Speed test; PV, Picture Vocabulary test.

### Partial Correlation Between CBS and Cognitive Measures

A partial correlation analysis was conducted to examine the relationship between CBS and cognitive measures while controlling for covariates. Detailed correlations can be found in Table 3 and Figure 2B. When controlling for age as a single covariate, a significant negative correlation between CBS scores and specific cognitive assessments, including MoCA (p = 0.017), DCCS (p < 0.001), and FICA (p = 0.039) tests, was identified. Remarkably, among these, only cognitive flexibility, as assessed by the DCCS test, displayed a robust negative correlation (see Table 3 for details). Conversely, there were no statistically significant associations between CBS scores and the PCPS, PSM, or PV tests.

**Table 3.**
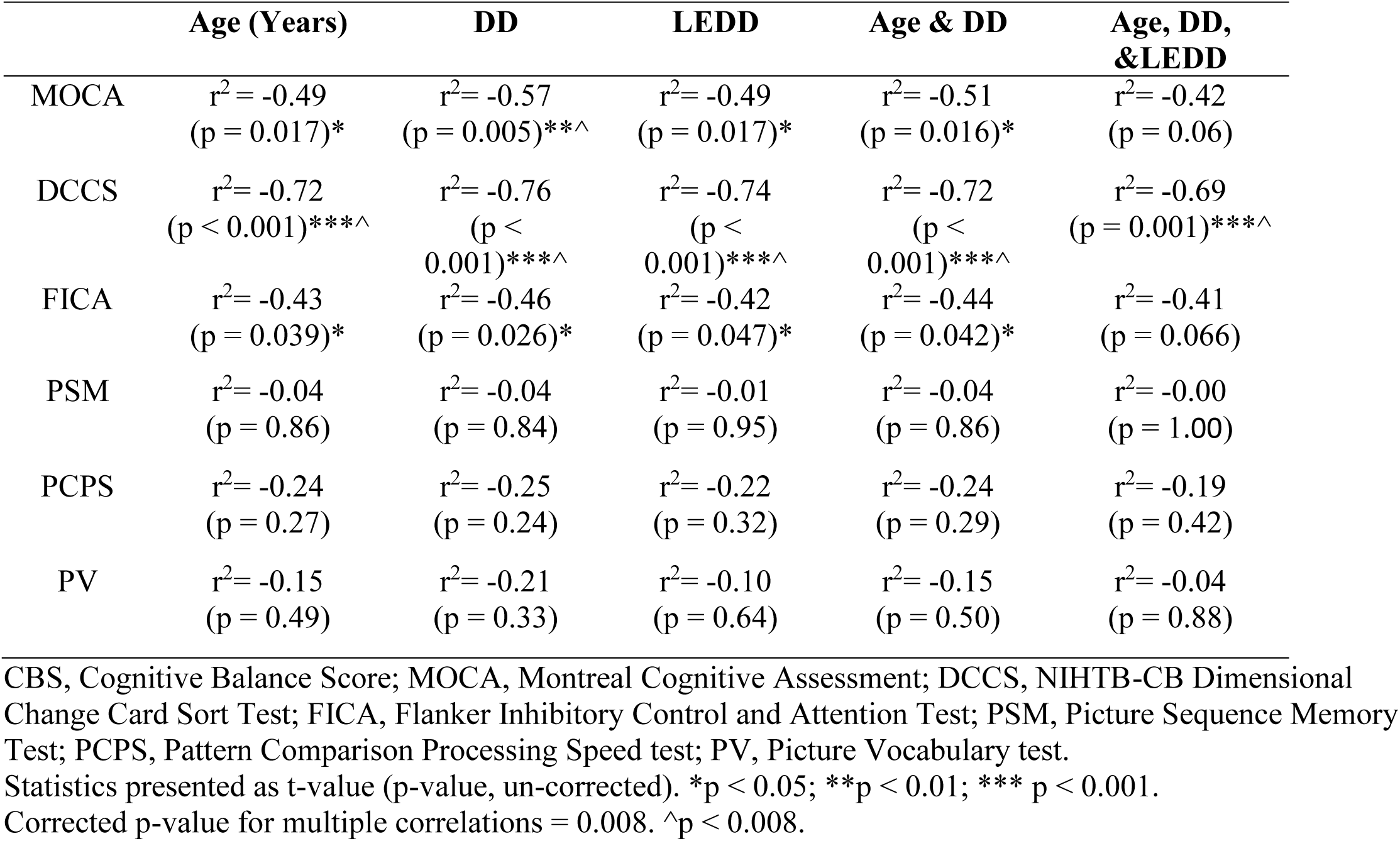
Pearson Partial Correlation between cognitive tests and CBS: Controlling for Age, Disease Duration (DD), and Levodopa Equivalent Daily Dose (LEDD).

When controlling for DD, a similar pattern emerged. There were strong negative correlations between CBS scores and the DCCS test (p < 0.001) and significant correlations between CBS scores, MoCA (p = 0.005), and FICA (p = 0.026) tests.

When LEDD was controlled, significant negative correlations between CBS scores and cognitive performance on the DCCS (p < 0.001) test were identified. Significant correlations were also present between CBS scores and MoCA (p = 0.017) and FICA (p = 0.047) assessments.

When simultaneously controlling for age and DD, several significant negative correlations persisted between CBS scores and measures of cognitive performance. The MoCA displayed a moderate negative correlation (p = 0.016) and the DCCS test exhibited a strong negative correlation (p < 0.001), suggesting a robust relationship between lower CBS scores and impaired cognitive flexibility. A significant negative correlation between CBS scores and the FICA test (p = 0.042) was also observed under this condition. Conversely, no statistically significant correlations occurred between CBS scores and the PCPS, PSM, or PV tests (p > 0.05).

Lastly, when controlling for all three covariates together (age, DD, and LEDD), only DCCS showed a significant negative correlation with CBS scores (p < 0.001). Under this condition, other cognitive domains did not display any significant associations.

When a Bonferroni-corrected p-value threshold of 0.008 was applied, a significant correlation between CBS scores and DCCS persisted. This correlation remained significant when controlling for age, DD, and LEDD individually, as well as in-combination. This finding represents a robust relationship between cognitive flexibility and clinical balance performance, regardless of which covariates are controlled for.

## DISCUSSION

### Cognitive Domains and Postural Control

The results of this investigation provide valuable insights into the complex relationship between cognitive domains and postural control in people with PD. This study employed a comprehensive measure of cognitive function, using the MoCA and the battery of cognitive tests included in the NIHTB-CB to evaluate various cognitive domains, including executive function, inhibitory control, processing speed, episodic memory, and language. We found that cognitive flexibility, a core component of executive functioning, exhibited a robust association with postural control. Specifically, individuals with PD who demonstrated impaired cognitive flexibility were more likely to experience postural instability. These results align with previous research that has highlighted the significance of executive function, including cognitive flexibility, in postural control (Fernandeset al, 2016; Kellyet al, 2015; Morriset al, 2019).

While cognitive flexibility emerged as a crucial factor, it is important to acknowledge that other cognitive domains also play roles in postural control for this population. However, after making corrections and controlling for covariates, our study did not observe statistically significant correlations between postural control and domains such as inhibitory control, processing speed, episodic memory, or language. Given the relatively small sample size (n=12 per group), these results should be interpreted cautiously, as limited statistical power may have contributed to these findings. This does not discount the potential contributions of other domains, as the multifaceted nature of postural control likely involves interactions between various cognitive processes (Johanssonet al, 2019; Kellyet al, 2015; Landerset al, 2005; Santamatoet al, 2015), but points towards a need for further research to explore these relationships more comprehensively.

It is also worth noting that postural stability outcomes are independent of gait outcomes in people with PD, implying that both posture and gait represent distinct facets of mobility.(Horak et al, 2016) It has been suggested that deficits associated with posture in PD can extend beyond cognitive processing and can encompass factors like cholinergic deficiency, abnormal body rigidity, leg muscle recruitment patterns, and body sway (Muller et al, 2013; Schoneburg et al, 2013). As a result of these complexities, the precise nature of the underlying network dysfunction responsible for postural instability in PD remains elusive.

Furthermore, previous research has shown that disorders characterized by executive dysfunction, such as ASD, OCD, MDD, and anorexia nervosa, can also exhibit impairments in cognitive flexibility (Miles et al, 2022; Sternheim et al, 2022). However, whether these impairments are directly linked to postural instability remains less clear. While some studies suggest altered balance control in ASD and OCD due to sensory integration deficits (Doumas et al, 2016; Oster and Zhou, 2022), the relationship between cognitive inflexibility and postural control in other conditions such as MDD and anorexia nervosa is largely unknown. Future research should explore whether similar relationships between cognitive flexibility and postural instability exist in these disorders characterized by executive dysfunction. If these populations do not exhibit similar postural deficits, this may suggest that postural instability in PD may be driven by broader cognitive-motor integration impairments rather than cognitive inflexibility particularly.

Given that our data also indicates significant correlations between MoCA scores and postural control, it is possible that the relationship observed in PD reflects a more global cognitive-motor integration impairment rather than cognitive flexibility specifically. The basal ganglia and cerebellar dysfunction in PD likely contribute to deficits in both cognitive flexibility and motor planning (Park et al, 2015; Yoshida et al, 2022), supporting this broader interpretation. Future studies should examine whether cognitive flexibility is uniquely predictive of postural instability across different neurological and psychiatric populations or if these associations are primarily driven by overarching deficits in cognitive-motor coordination.

### Potential Underlying Mechanisms

The observed relationship between cognitive flexibility and postural control in PD is likely mediated by dysfunction within the frontostriatal and cerebellar networks. The frontostriatal circuit, particularly involving the dorsolateral prefrontal cortex, basal ganglia, and thalamus, plays a crucial role in executive function, including cognitive flexibility (Basaia et al, 2022; Boschet al, 2021; Tait et al, 2025). The basal ganglia-thalamo-cortical loops, which regulate both motor planning and cognitive set-shifting, are known to be impaired in PD and may underlie the strong association between cognitive flexibility deficits and postural instability.

Additionally, the cerebellum, traditionally associated with motor coordination, has been increasingly recognized for its role in cognitive-motor integration. The cerebello-thalamo-cortical pathway, which connects the cerebellum to prefrontal regions, is critical for adapting motor responses based on cognitive demands (Aumann, 2002). Dysfunction in this network could contribute to impaired postural control in individuals with PD who exhibit cognitive inflexibility. These findings align with recent neuroimaging studies demonstrating reduced connectivity between the prefrontal cortex, basal ganglia, and cerebellum in PD patients with postural instability (Bagarinao et al, 2022). Given this evidence, our results suggest that interventions targeting these circuits, such as dual-task training or non-invasive neuromodulation of the prefrontal cortex and cerebellum, may hold promise for improving both cognitive and postural function in PD.

### Limitations

Several limitations of this study should be taken into consideration. First, the relatively small sample size may restrict the generalizability of findings to a broader population. A larger sample size would enhance statistical power and enable the detection of subtler relationships. Secondly, we acknowledge the limitation of not using a widely accepted, comprehensive assessment of postural stability, such as the BESTest or Mini BESTest, in our study. While CBS has been utilized in previous research (Boschet al, 2021; Pereraet al, 2018), it may not fully capture the complexity of postural control. Future studies should consider incorporating more detailed measures to provide a more comprehensive assessment of postural stability in PD. Thirdly, the cross-sectional design provides just a snapshot view of the role of cognitive flexibility in postural control; longitudinal studies could shed light on how these associations evolve with disease progression. Fourthly, the study predominantly focused on cognitive flexibility, but a more comprehensive examination of other cognitive domains, such as attention, memory, and visuospatial abilities, would provide a more holistic understanding of their contributions to postural control. Furthermore, there are limitations in linking cognition to broad constructs such as PIGD + and PIGD – or our study groups (PDPI+ and PDPI–), but this concern is mitigated by their established clinical relevance and the corroborative evidence from a continuous measure of balance, which aligns with the association between cognitive function and motor subtypes. Fifthly, excluding wheelchair-dependent PD patients with severe postural instability may introduce selection bias, potentially underrepresenting the most severely affected individuals. Lastly, the correlations outcomes establish associations but not causality, warranting further investigation into whether enhancing cognitive flexibility leads to improved postural control. These considerations underscore the complexity of the relationship between cognitive flexibility and postural stability in people with PD.

## CONCLUSION

This study underscores the critical role of cognitive flexibility in postural control among individuals with PD. The close relationship between cognitive flexibility deficits and postural instability highlights the significance of recognizing and addressing these cognitive impairments in clinical settings. This research adds depth to comprehension of the multifaceted nature of postural control in PD, opening opportunities for future investigations and interventions to enhance the well-being and safety of individuals who experience this symptom.

Additional opportunities for research might include directly examining neuromuscular responses during postural control tasks and conducting subgroup analyses within the PD group, based on factors like disease subtype and medication status. Our findings also suggest that cognitive flexibility should be considered when designing rehabilitation strategies. Given the strong association between cognitive flexibility and postural control, dual-task training—a motor-cognitive intervention that challenges patients to perform cognitive and motor tasks simultaneously—may be particularly beneficial. Recent studies have demonstrated that dual-task training can improve gait, balance, and executive function in PD (García-López et al, 2023; Johansson et al, 2023; Xiao et al, 2023). Therefore, integrating dual-task approaches into postural rehabilitation programs may enhance both cognitive and motor outcomes, ultimately improving functional mobility and reducing fall risk in individuals with PD. These interventions could encompass cognitive training regimens, dual-task training involving both cognitive and motor challenges, and strategies aimed at fortifying attention and cognitive flexibility during postural control exercises.

Further research should explore the causal links between cognitive flexibility and postural control while also focusing on the formulation of precise, targeted interventions aimed at optimizing outcomes for people with PD. The results of this study underscore the need for a comprehensive assessment of cognitive functions, with a particular focus on cognitive flexibility, in the clinical management of individuals with PD and postural instability.

## Data Availability

The datasets generated during and/or analyzed during the current study are available from the corresponding author on reasonable request.

## Abbreviations

PD: Parkinson’s disease
PDPI+: PD with postural instability
PDPI–: PD without postural instability
CBS: clinical balance score
MoCA: Montreal Cognitive Assessment
DCCS: Dimensional Change Card Sort Test
FICA: Flanker Inhibitory Control and Attention Test
PSM: Picture Sequence Memory Test
PCPS: Pattern Comparison Processing Speed test
PV: Picture Vocabulary test.

## REFERENCES

Amboni M, Barone P, Hausdorff JM. 2013. Cognitive contributions to gait and falls: Evidence and implications. Mov Disord. 28:1520–1533. doi:10.1002/mds.25674

Aumann TD. 2002. Cerebello-thalamic synapses and motor adaptation. Cerebellum. 1:69–77. doi:10.1080/147342202753203104

Bagarinao E, Kawabata K, Watanabe H, et al, 2022. Connectivity impairment of cerebellar and sensorimotor connector hubs in Parkinson’s disease. Brain Communications. 4. doi:10.1093/braincomms/fcac214

Barboza NM, Mancini M, Smaili SM, et al, 2023. Exploring mobility dysfunction in people with and without impaired cognition in Parkinson disease. Parkinsonism Relat Disord. 115:105836. doi:10.1016/j.parkreldis.2023.105836

Basaia S, Agosta F, Francia A, et al, 2022. Cerebro-cerebellar motor networks in clinical subtypes of Parkinson’s disease. npj Parkinson’s Disease. 8:113. doi:10.1038/s41531-022-00377-w

Bosch TJ, Kammermeier S, Groth C, et al, 2021. Cortical and Cerebellar Oscillatory Responses to Postural Instability in Parkinson’s Disease. Front Neurol. 12:752271. doi:10.3389/fneur.2021.752271

Buated W, Lolekha P, Hidaka S, et al, 2016. Impact of Cognitive Loading on Postural Control in Parkinson’s Disease With Freezing of Gait. Gerontol Geriatr Med. 2:2333721416673751. doi:10.1177/2333721416673751

Chaudhary S, Kumaran SS, Kaloiya GS, et al, 2020. Domain specific cognitive impairment in Parkinson’s patients with mild cognitive impairment. J Clin Neurosci. 75:99–105. doi:10.1016/j.jocn.2020.03.015

Dajani DR, Uddin LQ. 2015. Demystifying cognitive flexibility: Implications for clinical and developmental neuroscience. Trends Neurosci. 38:571–578. doi:10.1016/j.tins.2015.07.003

Diamond A. 2013. Executive functions. Annu Rev Psychol. 64:135–168. doi:10.1146/annurev-psych-113011-143750

Doumas M, McKenna R, Murphy B. 2016. Postural Control Deficits in Autism Spectrum Disorder: The Role of Sensory Integration. J Autism Dev Disord. 46:853–861. doi:10.1007/s10803-015-2621-4

Fernandes A, Mendes A, Rocha N, et al, 2016. Cognitive predictors of balance in Parkinson’s disease. Somatosens Mot Res. 33:67–71. doi:10.1080/08990220.2016.1178634

Frenklach A, Louie S, Koop MM, et al, 2009. Excessive postural sway and the risk of falls at different stages of Parkinson’s disease. Mov Disord. 24:377–385. doi:10.1002/mds.22358

García-López H, de los Ángeles Castillo-Pintor M, Castro-Sánchez AM, et al, 2023. Efficacy of Dual-Task Training in Patients with Parkinson’s Disease: A Systematic Review with Meta-Analysis. Movement Disorders Clinical Practice. 10:1268–1284. doi:10.1002/mdc3.13823

Green J, McDonald W, Vitek J, et al, 2002. Cognitive impairments in advanced PD without dementia. Neurology. 59:1320–1324.

Hanes KR, Andrewes DG, Pantelis C. 1995. Cognitive flexibility and complex integration in Parkinson’s disease, Huntington’s disease, and schizophrenia. J Int Neuropsychol Soc. 1:545–553. doi:10.1017/s1355617700000679

Horak FB, Mancini M, Carlson-Kuhta P, et al, 2016. Balance and Gait Represent Independent Domains of Mobility in Parkinson Disease. Phys Ther. 96:1364–1371. doi:10.2522/ptj.20150580

Johansson H, Folkerts AK, Hammarström I, et al, 2023. Effects of motor-cognitive training on dual-task performance in people with Parkinson’s disease: a systematic review and meta-analysis. J Neurol. 270:2890–2907. doi:10.1007/s00415-023-11610-8

Johansson H, Franzen E, Skavberg Roaldsen K, et al, 2019. Controlling the Uncontrollable: Perceptions of Balance in People With Parkinson Disease. Phys Ther. 99:1501–1510. doi:10.1093/ptj/pzz117

Kelly VE, Johnson CO, McGough EL, et al, 2015. Association of cognitive domains with postural instability/gait disturbance in Parkinson’s disease. Parkinsonism Relat Disord. 21:692–697. doi:10.1016/j.parkreldis.2015.04.002

Landers M, Wulf G, Wallmann H, et al, 2005. An external focus of attention attenuates balance impairment in patients with Parkinson’s disease who have a fall history. Physiotherapy. 91:152–158. doi:10.1016/j.physio.2004.11.010

Lyros E, Messinis L, Papathanasopoulos P. 2008. Does motor subtype influence neurocognitive performance in Parkinson’s disease without dementia? Eur J Neurol. 15:262–267.

Miles S, Phillipou A, Sumner P, et al, 2022. Cognitive flexibility and the risk of anorexia nervosa: An investigation using self-report and neurocognitive assessments. J Psychiatr Res. 151:531–538. doi:10.1016/j.jpsychires.2022.05.043

Morris R, Martini DN, Smulders K, et al, 2019. Cognitive associations with comprehensive gait and static balance measures in Parkinson’s disease. Parkinsonism Relat Disord. 69:104–110. doi:10.1016/j.parkreldis.2019.06.014

Muller ML, Albin RL, Kotagal V, et al, 2013. Thalamic cholinergic innervation and postural sensory integration function in Parkinson’s disease. Brain. 136:3282–3289. doi:10.1093/brain/awt247

Nasreddine ZS, Phillips NA, Bedirian V, et al, 2005. The Montreal Cognitive Assessment, MoCA: a brief screening tool for mild cognitive impairment. J Am Geriatr Soc. 53:695–699. doi:10.1111/j.1532-5415.2005.53221.x

Nutt JG, Horak FB, Bloem BR. 2011. Milestones in gait, balance, and falling. Mov Disord. 26:1166–1174. doi:10.1002/mds.23588

Nyholm D, Jost WH. 2021. An updated calculator for determining levodopa-equivalent dose. Neurol Res Pract. 3:58. doi:10.1186/s42466-021-00157-6

Oster LM, Zhou G. 2022. Balance and Vestibular Deficits in Pediatric Patients with Autism Spectrum Disorder: An Underappreciated Clinical Aspect. Autism Research and Treatment. 2022:7568572. doi:10.1155/2022/7568572

Pal G, O’Keefe J, Robertson-Dick E, et al, 2016. Global cognitive function and processing speed are associated with gait and balance dysfunction in Parkinson’s disease. J Neuroeng Rehabil. 13:94. doi:10.1186/s12984-016-0205-y

Park JH, Kang YJ, Horak FB. 2015. What Is Wrong with Balance in Parkinson’s Disease? J Mov Disord. 8:109–114. doi:10.14802/jmd.15018

Pelicioni PHS, Menant JC, Latt MD, et al, 2019. Falls in Parkinson’s Disease Subtypes: Risk Factors, Locations and Circumstances. Int J Environ Res Public Health. 16. doi:10.3390/ijerph16122216

Perera T, Tan JL, Cole MH, et al, 2018. Balance control systems in Parkinson’s disease and the impact of pedunculopontine area stimulation. Brain. 141:3009–3022. doi:10.1093/brain/awy216

Santamato A, Ranieri M, Cinone N, et al, 2015. Postural and Balance Disorders in Patients with Parkinson’s Disease: A Prospective Open-Label Feasibility Study with Two Months of Action Observation Treatment. Parkinsons Dis. 2015:902738. doi:10.1155/2015/902738

Schoneburg B, Mancini M, Horak F, et al, 2013. Framework for understanding balance dysfunction in Parkinson’s disease. Mov Disord. 28:1474–1482. doi:10.1002/mds.25613

Sousa NMF, Macedo RC, Brucki SMD. 2021. Cross-sectional associations between cognition and mobility in Parkinson’s disease. Dement Neuropsychol. 15:105–111. doi:10.1590/1980-57642021dn15-010011

Sternheim LC, van Passel B, Dingemans A, et al, 2022. Cognitive and Experienced Flexibility in Patients With Anorexia Nervosa and Obsessive Compulsive Disorder. Front Psychiatry. 13:868921. doi:10.3389/fpsyt.2022.868921

Tait P, Graham L, Vitorio R, et al, 2025. Neuroimaging and cognitive correlates of postural control in Parkinson’s disease: a systematic review. J Neuroeng Rehabil. 22:24. doi:10.1186/s12984-024-01539-y

Weintraub S, Dikmen SS, Heaton RK, et al, 2013. Cognition assessment using the NIH Toolbox. Neurology. 80:S54–64. doi:10.1212/WNL.0b013e3182872ded

Xiao Y, Yang T, Shang H. 2023. The Impact of Motor-Cognitive Dual-Task Training on Physical and Cognitive Functions in Parkinson’s Disease. Brain Sciences. 13:437.

Yoshida J, Oñate M, Khatami L, et al, 2022. Cerebellar Contributions to the Basal Ganglia Influence Motor Coordination, Reward Processing, and Movement Vigor. J Neurosci. 42:8406–8415. doi:10.1523/jneurosci.1535-22.2022

